# Input of Major Histocompatibility Complex class II immunostaining in idiopathic inflammatory myopathies

**DOI:** 10.1101/2022.11.28.22282671

**Authors:** Lola E. R. Lessard, Marie Robert, Tanguy Fenouil, Véréna Landel, Marie Carlesimo, Arnaud Hot, Bénédicte Chazaud, Thomas Laumonier, Nathalie Streichenberger, Laure Gallay

## Abstract

**Background:** Idiopathic inflammatory myopathies (IIMs) are a group of rare acquired muscular diseases. In healthy muscle, myofibers do not express major histocompatibility complex (MHC) class I and II. It was established that MHC-I positive immunostaining, although non-specific, is a marker for IIM diagnosis, while the significance of MHC-II immunostaining remains unclear. The present study investigates the expression of MCH-II in myofibers and capillaries of IIM muscles, taking into account the current IIM classification.

**Patients & Methods:** A historical cohort was designed, including dermatomyositis (DM), inclusion body myositis (IBM), anti-synthetase syndrome (ASyS), immune-mediated necrotizing myopathy (IMNM), or overlap myositis (OM). MHC-II immunostaining was performed on patient muscle sections and was analyzed in a standardized and blind manner.

**Results:** Muscle sections from biopsies of 72 IIM patients were included: 23 DM, 17 IBM, 12 IMNM, 9 ASyS, and 11 OM. Overall, abnormal MHC-II immunostaining was found in myofibers and/or capillaries in 67 (93%) patients. Myofiber MHC-II immunostaining patterns differed according to the IIM subgroup: the immunostaining was diffuse in IBM (100%), negative in IMNM (75%), perifascicular in ASyS (67%), and either diffuse heterogeneous, clustered, or perifascicular in OM (27%, 27%, and 18%, respectively). MHC-II expression was found in 50% of DM (n=11/22).

While all IIM subgroups presented quantitative and qualitative abnormalities of MHC-II immunostaining in capillaries, some subgroups displayed specificities. Most IBM and IMNM muscles presented frequent dilated capillaries (88% and 67%, respectively). DM, ASyS, and OM exhibited high frequencies of capillary lesions, including capillary dropout, leaky capillaries, and dilated capillaries.

**Conclusion:** While recent expert opinion (EURO-NMD pathology working group) recommended that MHC-II immunostaining of muscle biopsy remains optional, the present work demonstrates that the expression pattern of MHC-II allows to distinguish between several IIM subgroups. Our data argue for the inclusion of MHC-II immunostaining in the routine histological diagnosis for IIMs.

## Introduction

Idiopathic inflammatory myopathies (IIMs) are rare acquired muscle disorders. In the last decade, the increasing understanding of IIMs has allowed the identification of five main clinico-pathological subgroups: dermatomyositis (DM), inclusion body myositis (IBM), immuno-mediated necrotizing myopathy (IMNM), anti-synthetase syndrome (ASyS), and overlap myositis (OM) [2, 22, 28, 31, 32, 49]. These IIM subgroups differ in terms of clinical features, muscle biopsy analysis, and auto-antibodies detected, notably myositis associated antibodies (MSA). In clinical practice, the final diagnosis of IIM is sometimes difficult to reach, which can lead to a significant delay in patient care [35]. Since therapeutic approaches are highly variable among IIM subgroups, reducing diagnostic delay could significantly improve patient management.

The major histocompatibility complex (MHC) corresponds to cell surface glycoproteins entrusted with antigen presentation. The two major classes of MHC, MHC-I and MHC-II molecules, present antigenic peptides to CD8+ T cells and CD4+ T cells, respectively [40]. All nucleated cells express MHC-I while MHC-II is normally expressed only by “professional” antigen-presenting cells (APCs), and can be expressed by several other cell types upon inflammatory signals [42]. Histological analysis shows that healthy and mature human skeletal muscle fibers do not express MHC-I nor MHC-II, while endothelial cells, *i*.*e*. capillaries, present a punctiform positive immunostaining for both MHCs [26, 44, 53]. Myofiber expression of MHC-I, also known as HLA-ABC, is considered as a diagnostic tool for IIMs, with high sensitivity but low specificity [9, 14, 24, 36, 39, 43]. Few studies have investigated the expression of MHC-II (also known as HLA-DR/HLA-DQ/HLA-DP) in IIM muscles [4, 6, 14, 17, 23, 24, 33, 39, 43, 54]. To our knowledge, except for one study specific to ASyS [4], the other studies were realized from 10 to 50 years ago, and thus did not take into account the current IIM classification. Consequently, recent expert opinion (EURO-NMD pathology working group) recommended that MHC-II immunostaining of muscle biopsy remains optional for the diagnosis of IIMs [51]. Furthermore, several studies demonstrated that MHC-II immunostaining distinguishes IIMs and hereditary myopathies (positivity in 61.7% and 10.1%, respectively), while MHC-I positivity was frequent in both groups (98.3% and 92.7%, respectively) [10, 23, 43], highlighting the sensitivity of MHC-I immunostaining and the specificity of MHC-II immunostaining, thereby strengthening the usefulness of MHC-II immunostaining in clinical practice.

In the present study, the expression of MCH-II in myofibers and capillaries of IIM muscles was investigated, taking into account the current IIM classification, with the aim to evaluate the usefulness of MCH-II immunostaining for the diagnosis of IIM.

## Materials & Methods

### Patients

A historical cohort was created using the MYOLYON register by screening for patients diagnosed with IIM between 2017 and 2021. The inclusion criterion was a final diagnosis of DM, IBM, ASyS, IMNM, or OM in accordance with the current international IIM diagnosis [2, 28, 29, 31]. All patients had a muscle biopsy (in deltoid or vastus lateralis muscle) and had benefited from the recommended neuropathological analysis.

### Histopathology

All the immunostainings were performed with an immunohistochemistry automated instrument (Ventana, Ultra Benchmark, Ventana Medical Systems, Inc., Tucson, Arizona, USA) on 7 μm muscle cryosections using the following primary antibodies: anti-MHC-I (clone W6/32, DAKO, ref: M0736, 1/2400), anti-MHC-II (clone CR3/43, DAKO, ref: M0775, 1/400, corresponding to Anti-HLA DR + DP + DQ antibody), anti-CD56 (clone 123C3, Cell Marque, ref:156-M85, 1/50) Leica Biosystems NCL-CD56-1B6), and anti-CD31 (clone JC70A, Ventana, ref: 760-4378, prediluted), anti-slow myosin heavy chain (Novocastra, ref: NCL-MHCs, 1/80) and anti-fast myosin heavy chain (Novocastra, ref: NCL-MHCf, 1/40). All primary antibodies were revealed using the ultraView Universal DAB Detection Kit (Ventana, ref: 05269806001) except for the anti-slow myosin heavy chain that was revealed using the ultraView Universal AP Red Detection Kit (Ventana, ref: 05269814001). Before sMHC and fMHC antibody revelations, an amplification step was added using OptiView Amplification Kit, Ventana, ref: 760-099) (Online Resource 1). For each staining, a healthy control muscle sample was added on each slide. Immunostainings were analyzed in a blind manner. The positivity of immunostaining was evaluated in a minimum of three representative fascicles, and defined in the most affected fascicle by (i) the presence of positive myofibers in the whole fascicle, and (ii) the distribution of the positive myofibers. The expression of MHC-II and MHC-I by myofibers was considered positive when sarcolemmal and/or sarcoplasmic staining was observed. Biopsies with questionable or faint staining or fainter than the background (i.e. healthy muscle control present on each slide) were considered as negative. Immunostainings were evaluated on serial sections of the same sample, allowing the evaluation of the co-expression of MHC-I, MHC-II, and CD56 (expressed by regenerating myofibers) by the same myofibers, and by CD31 positive structures (gold standard for capillary endothelial cell immunostaining) [51]. CD56 positivity was defined by a sarcoplasmic immunostaining in at least 5 myofibers per fascicle. Digital image capture was performed using an Axio Scan.Z1® (Zeiss).

### Statistical analysis

Descriptive analysis and frequency calculations were performed using Microsoft Excel. Medians were reported with interquartile range [IQR].

## Results

A total of 72 patients with a final diagnosis of IIM and available muscle biopsies were included: 23 DM, 17 IBM, 12 IMNM, 9 ASyS, and 11 OM. The overall median [IQR] age was 60 years old [44-70] and sex ratio was 0.8. Among the 72 patients, 5 received immunosuppressive drugs before the muscle biopsy (ASyS n=2, DM, IBM, OM n=1 each). No specific MSA was preferentially detected in DM patients (Online Resource 2). Regarding other patients, 7/17 IBM presented anti-Cn1A, 9/12 IMNM had either anti-HMGCoA antibodies (Ab) (n=8) or anti-SRP Ab (n=1), ASyS patients all had anti-RNA synthetase Ab (anti Jo1 n=6, anti PL7 n=3), and 6/11 OM patients had auto-antibodies (anti-Ku n=2, anti Scl70 n=2, anti SmRNP n=2). Overall, abnormal MHC-II immunostaining was found in myofibers and/or capillaries in 67 (93%) patients: 22 (96%) DM, 17 (100%) IBM, 9 (75%) IMNM, 8 (89%) ASyS, and 11 (100%) OM patients.

### MYOFIBER IMMUNOHISTOCHEMISTRY STUDY

#### Abnormal expression of MHC-II, MHC-I, and CD56 in IIM myofibers

Myofiber MHC-II immunostaining exhibited 5 different patterns: negative, diffuse, perifascicular, scattered, or clustered. Diffuse positive immunostaining was either homogenous or heterogenous. Perifascicular immunostaining was either strictly perifascicular or extended perifascicular (Figure 1, Table 1). Overall, myofibers showed a positive expression of MHC-II in 47 (65%) patients. The variations in MHC-II myofiber patterns were as follows: negative (n=24/72, 33%), diffuse homogenous (n=5/72, 7%), diffuse heterogenous (16/72, 22%), strictly perifascicular (n=15/72, 21%), extended perifascicular (n=3/72, 4%), scattered (n=4/72, 6%), and clustered (n=7/72, 10%) (Table 2).

**Table 1.** Definitions of the various immunostaining patterns.

| **Myofiber MHC-II and MHC-I immunostaining patterns** |  |  |
| --- | --- | --- |
| Diffuse |  |  |
| Homogenous |  | Diffuse and homogenous myofiber positivity in the muscle biopsy |
| Heterogenous |  | Diffuse and heterogenous myofiber positivity in the muscle biopsy |
| Perifascicular pattern |  |  |
| Strictly perifascicular |  | Positive myofibers only in the perifascicular area, extended to a maximum of 4-cell layers |
| Extended perifascicular |  | Diffuse positive immunostaining of myofibers according to a positivity gradient with a maximal intensity in the perifascicular area |
| Scattered |  | Positive myofibers scattered throughout the muscle biopsy with at least 5 myofibers per fascicle |
| Clustered* |  | Positive immunostaining of myofibers around necrotic myofibers and/or inflammatory infiltrates, including a minimum of 10 positive myofibers |
| Negative |  | No myofiber positivity |
| **Capillary MHC-II and CD31 immunostaining patterns** |  |  |
| Capillary dropout |  | At least 3 myofibers with no related capillaries, found at least 3 times in the muscle biopsy |
| Leaky |  | Positive immunostaining of capillaries brimming over capillary edges, giving a blurred aspect |
| Dilated |  | Positive immunostaining of oversize capillaries with marked thickening and diameter-increasing in transversal section |
| Normal |  | Punctiform immunostaining without visible lumen |
| MHC: major histocompatibility complex. *only found with MHC-II immunostaining. |  |  |

**Table 2.** Major histocompatibility complex (MHC) class II, I, and CD56 myofiber immunostaining in IIM patients.

|  | DM | IBM | IMNM | ASyS | OM | total |
| --- | --- | --- | --- | --- | --- | --- |
| Patients (n) | 23 | 17 | 12 | 9 | 11 | 72 |
| **MHC-II pattern** |  |  |  |  |  |  |
| MHC-II positive myofibers | 11/23<br>(48%) | 17/17<br>(100%) | 3/12<br>(25%) | 8/9<br>(89%) | 8/11<br>(73%) | 47/72<br>(65%) |
| Diffuse MHC-II expression | 1/23 (4%) | 16/17<br>(94%) | 0 | 2/9<br>(22%) | 3/11<br>(27%) | 24/72<br>(33%) |
| - homogenous | 1/23 (4%) | 4/17<br>(24%) | 0 | 0 | 0 | 5/72 (7%) |
| - heterogenous | 0 | 12/17<br>(71%) | 0 | 1/9<br>(11%) | 3/11<br>(27%) | 16/72<br>(22%) |
| Perifascicular MHC-II expression | 9/23<br>(39%) | 1/17 (6%) | 0 | 6/9<br>(67%) | 2/11<br>(18%) | 18/72<br>(25%) |
| - strictly perifascicular | 7/23<br>(30%) | 1/17 (6%) | 0 | 5/9<br>(56%) | 2/11<br>(18%) | 15/72<br>(21%) |
| - extended perifascicular | 2/23 (9%) | 0 | 0 | 1/9<br>(11%) | 0 | 3/72 (4%) |
| Scattered MCH-II expression | 0 | 0 | 3/12<br>(25%) | 0 | 1/11 (9%) | 4/72 (6%) |
| Cluster MHC-II expression | 2/23 (9%) | 0 | 0 (0%) | 2/9<br>(22%) | 3/11<br>(27%) | 7/72<br>(10%) |
| MHC-II negative myofibers | 12/23<br>(52%) | 0 | 9/12<br>(75%) | 1/9<br>(11%) | 3/11<br>(27%) | 24/72<br>(33%) |
| **MHC-I pattern** |  |  |  |  |  |  |
| MHC-I positive | 22/23<br>(96%) | 17/17<br>(100%) | 10/12<br>(83%) | 9/9<br>(100%) | 10/11<br>(91%) | 68/72<br>(94%) |
| Diffuse MHC-I expression | 10/23<br>(43%) | 17/17<br>(100%) | 2/12<br>(17%) | 2/9<br>(22%) | 6/11<br>(55%) | 55/72<br>(76%) |
| - homogenous | 10/23<br>(43%) | 10/17<br>(59%) | 0 | 1/9<br>(11%) | 5/11<br>(45%) | 26/72<br>(36%) |
| - heterogenous | 0 | 7/17<br>(41%) | 2/12<br>(17%) | 1/9<br>(11%) | 1/11 (9%) | 15/72<br>(21%) |
| Perifascicular MHC-I | 12/23<br>(52%) | 0 | 0 | 6/9<br>(67%) | 4/11<br>(36%) | 22/72<br>(31%) |
| - extended perifascicular | 8/23<br>(35%) | 0 | 0 | 4/9<br>(44%) | 3/11<br>(27%) | 15/72<br>(21%) |
| - strictly perifascicular | 4/23<br>(17%) | 0 | 0 | 2/9<br>(22%) | 1/11 (9%) | 7/72<br>(10%) |
| Scattered MHC-I expression | 0 | 0 | 8/12<br>(67%) | 1/9<br>(11%) | 0 | 6/72 (8%) |
| MHC-I negative myofibers | 1/23 (4%) | 0 | 2/12<br>(17%) | 0 | 1/11 (9%) | 4/72 (6%) |
| Co-staining with MHC-II | 4/23<br>(17%) | 17/17<br>(100%) | 3/12<br>(25%) | 5/9<br>(56%) | 4/11<br>(36%) | 33/72<br>(46%) |
| **CD56** |  |  |  |  |  |  |
| CD56 positive myofibers | 22/22<br>(100%) | 17/17<br>(100%) | 12/12<br>(100%) | 7/7<br>(100%) | 10/11<br>(91%) | 68/69<br>(99%) |
| Co-staining with MHC-II | 10/22<br>(45%) | 17/17<br>(100%) | 3/12<br>(25%) | 7/7<br>(100%) | 10/11<br>(91%) | 47/69<br>(68%) |
| Co-staining in MHC-II positive cases | 7/11<br>(64%) | 17/17<br>(100%) | 3/3<br>(100%) | 7/7<br>(100%) | 7/7<br>(100%) | 41/45<br>(91%) |

**Fig. 1.**
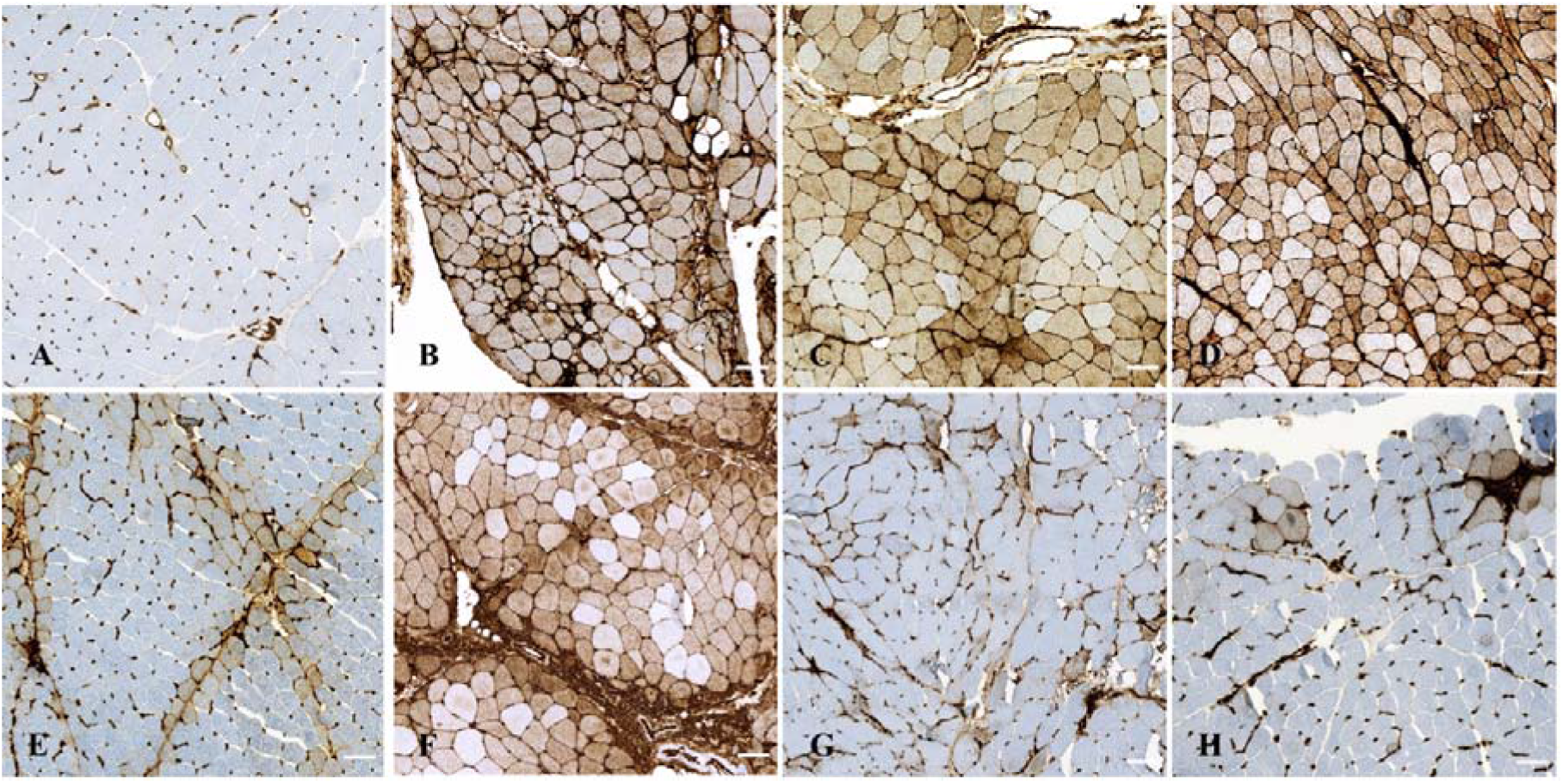
Myofiber major histocompatibility complex (MHC) class II immunostaining patterns in muscle biopsies from controls and IIM patients. Normal expression in control muscle biopsy (A). Diffuse homogenous myofiber positivity (B). Diffuse heterogeneous myofiber positivity (C and D). Strictly perifascicular myofiber positivity (E). Extended perifascicular myofiber positivity (F). Scattered myofiber positivity (G). Clustered myofiber positivity (H). Bars = 100μm.

MHC-I expression by myofibers was largely observed in IIM patients (n=68/72, 94%). MHC-I staining exhibited the same types of patterns except that positive clustered myofiber pattern was not found (Figure 2, Table 1). The various MHC-I positive myofiber patterns were found as follows: negative (n=4/72, 6%), diffuse homogenous (n=26/72, 36%), diffuse heterogenous (n=15/72, 21%), strictly perifascicular (n=7/72, 10%), extended perifascicular (n=15/72, 21%), and scattered (n=6/72, 8%) (Figure 2, Table 3).

**Table 3.** MHC-II and CD31 capillary immunostaining in IIM patients.

|  | DM | IBM | IMNM | ASyS | OM | total |
| --- | --- | --- | --- | --- | --- | --- |
| Patients (n) | 23 | 17 | 12 | 9 | 11 | 72 |
| **Capillary quantitative abnormality: capillary dropout** |  |  |  |  |  |  |
| MHC-II capillary dropout | 17/23<br>(74%) | **1/9**<br>**(11%)\*** | 2/12<br>(17%) | 4/9<br>(44%) | 8/11<br>(73%) | 32/64 (50%) |
| CD31 capillary dropout | **18/22**<br>**(82%)** | 1/16<br>(6%) | **6/11**<br>**(55%)** | **6/9**<br>**(67%)** | **8/9**<br>**(89%)** | 39/67 (58%) |
| *Correlation of CD31 with MHC-II* | 20/22<br>(91%) | 9/9<br>(100%) | 7/11<br>(64%) | 7/9<br>(78%) | 7/9<br>(78%) | 50/60 (83%) |
| **Capillary qualitative abnormalities: leaky or dilated capillaries** |  |  |  |  |  |  |
| MHC-II capillary structural abnormalities | 20/22<br>(91%) | 16/16<br>(100%) | 10/11<br>(91%) | 9/9<br>(100%) | 10/11<br>(91%) | 65/72 (90%) |
| Leaky MHC-II capillaries | 20/23<br>(87%) | **4/7**<br>**(57%)\*** | **8/12**<br>**(67%)** | **6/9**<br>**(67%)** | **10/11**<br>**(91%)** | 48/62 (77%) |
| Leaky CD31 capillaries | **21/22**<br>**(95%)** | 7/16<br>(44%) | 6/11<br>(55%) | 5/9<br>(56%) | 7/9<br>(78%) | 46/67 (69%) |
| *Correlation of CD31 with MHC-II* | 22/22<br>(100%) | 7/7<br>(100%) | 9/11<br>(82%) | 8/9<br>(89%) | 8/9<br>(89%) | 54/58 (93%) |
| Dilated MHC-II capillaries | 17/23<br>(74%) | 15/17<br>(88%) | 8/12<br>(67%) | 8/9<br>(89%) | 5/11<br>(45%) | 53/72 (74%) |
| Dilated CD31 capillaries | **19/22**<br>**(86%)** | **15/16**<br>**(94%)** | **8/11**<br>**(73%)** | **8/9**<br>**(89%)** | **7/9**<br>**(78%)** | 57/67 (85%) |
| *Correlation of CD31 with MHC-II* | 19/22<br>(86%) | 13/16<br>(81%) | 10/11<br>(91%) | 9/9<br>(100%) | 7/9<br>(78%) | 58/67 (87%) |

In bold the highest value of an elementary lesion (i.e. capillary dropout, leaky, or dilated capillaries), in red the highest value for each IIM subgroup. \* difficult to assess due to diffuse myofiber positivity.

**Fig. 2.**
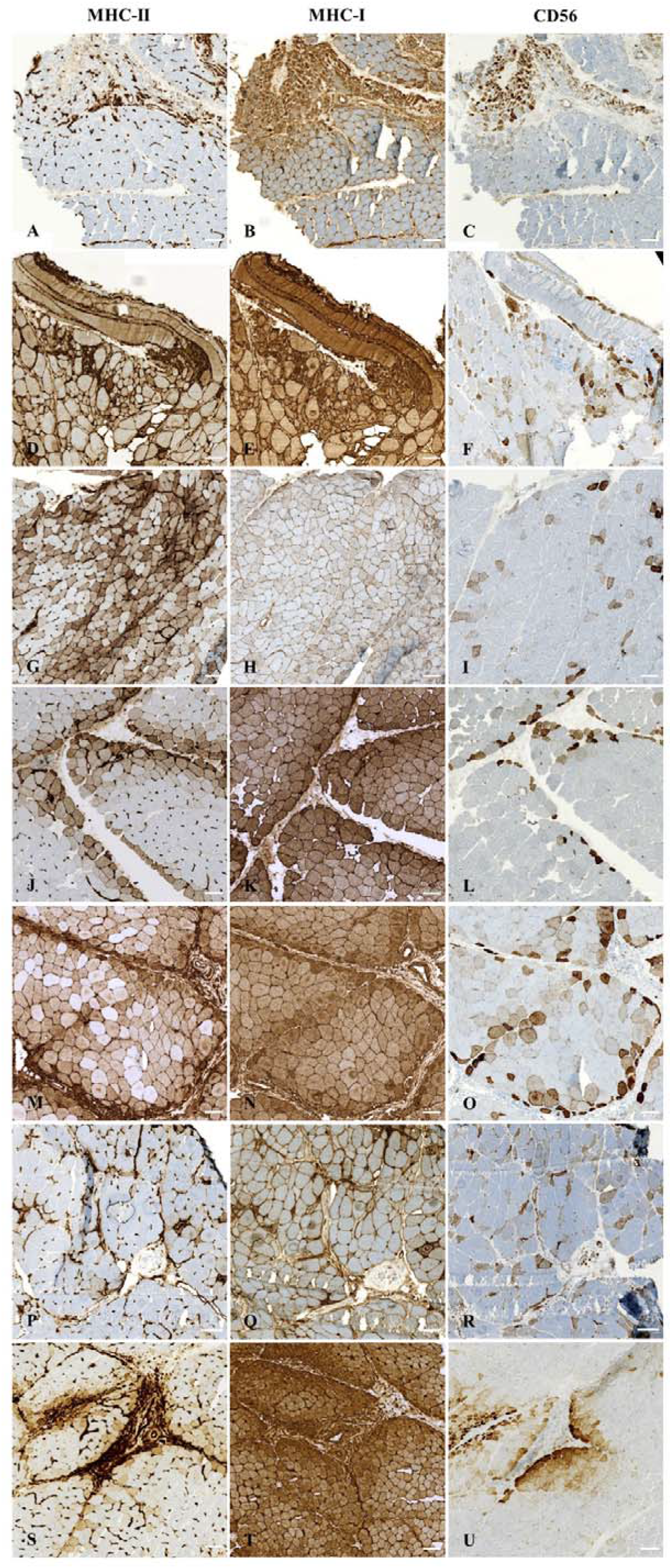
Major histocompatibility complex (MHC) class I and II and CD56 myofiber immunostaining in illustrative muscle biopsies from 8 IIM patients. Left panel shows MHC-II immunostaining, middle panel shows MHCI-I immunostaining and right panel show CD56 immunostaining on serial sections of 7 IIM patients. (A-C) negative MHC-II myofiber immunostaining (A), and corresponding diffuse MHC-I (B) and CD56 (C) immunostaining. (D-F) diffuse homogenous MHC-II myofiber positivity (D), and corresponding diffuse MHC-I (E) and CD56 (F) immunostaining. (G-I) diffuse heterogeneous MHC-II myofiber positivity (G) and corresponding diffuse MHC-I (H) and CD56 (I) immunostaining. (J-L) strictly perifascicular MHC-II myofiber positivity (K) and corresponding diffuse MHC-I (L) and CD56 (M) immunostaining. (M-O) Extended perifascicular MHC-II myofiber positivity (M) and corresponding diffuse MHC-I (N) and CD56 (O) immunostaining. (P-R) scattered MHC-II myofiber positivity (P) and corresponding diffuse MHC-I (Q) and CD56 (R) immunostaining. (S-U) clustered MHC-II myofiber positivity (S) and corresponding diffuse MHC-I (T) and CD56 (U) immunostaining. Bars = 100μm.

CD56 was expressed by some IIM myofibers in all but one tested muscle biopsies (n=68/69, 99%; Figure 2).

#### Differential myofiber MHC-II expression according to IIM subgroup

The MHC-II immunostaining patterns differed according to the IIM subgroup (Figure 2, Table 2). IBM muscles showed MHC-II positive myofibers in 100% of patients, mainly with diffuse immunostaining (n=16/17, 94%). IMNM patients presented mainly negative MHC-II myofiber immunostaining (n=9/12, 75%), and the 3 patients with positive MHC-II immunostaining presented only scattered myofiber positivity. Myofiber MHC-II expression was positive in 89% of the AsyS patients, with mainly perifascicular MHC-II immunostaining (n=6/9, 67%). Among OM patients, 73% exhibited with MHC-II myofiber expression, with either diffuse heterogenous (n=3/11, 27%), clustered (n=3/11, 27%), or perifascicular (n=2/11, 18%) MHC-II positivity. DM muscles showed an inconsistent expression of MHC-II by myofibers. In 52% (12/23) of the muscle biopsies, myofibers did not express MHC-II, while MHC-II positive myofibers were observed as either diffuse homogenous (1/23, 4%), perifascicular (n=9/23, 39%), or clustered (2/23, 9%, one having both perifascicular and clustered positivity). Given this variability in MHC-II immunostaining, we decided to further investigate DM cases. Among the DM patients with MHC-II positive myofibers, no specific MSA was preferentially detected. Nevertheless, a substantial number of these DM patients were juvenile cases (5/11, 45%) or displayed ongoing neoplasia (n=4/11, 36%). Conversely, among all DM patients (n=23), 6 had paraneoplastic DM; the 4 (67%) who presented MHC-II positive myofibers had lung adenocarcinoma (n=2) or melanoma (n=2) while the other 2 (33%) had ovarian adenocarcinoma. In the present study, 7 juvenile DM cases were included; the 5 patients (71%) who had MHC-II positive myofiber immunostaining were all aged under 10 years old (0-5 years: 2 patients, 6-10 years: 3 patients), while the other 2 (29%) were between 11 and 15 years old (Online Resource 2).)

#### Differential MHC-I expression by myofibers according to IIM subgroup

Myofiber MHC-I expression differed according to IIM subgroups (Table 2). All IBM patients presented a diffuse MHC-I immunostaining, while in IMNM, a majority of patients displayed a scattered myofiber positive immunostaining (8/12, 67%). In ASyS, myofiber MHC-I immunostaining was mainly perifascicular (6/9, 67%) with either extended perifascicular (4/9, 44%) or strictly perifascicular (2/9, 22%) positivity. Myofiber MHC-I expression was positive in OM (10/11, 91%), with mainly a diffuse pattern of positivity (n=6/11, 55%) or a perifascicular pattern (n=4/11, 36%). Finally, DM patients presented diffuse homogenous myofiber MHC-I immunostaining (n=10/23, 43%) or a perifascicular pattern (n=12/23, 52%). In addition, there was no obvious co-expression of MHC-I and MHC-II in DM, IMNM, and OM while in ASyS muscles both MHC-I and MHC-II immunostaining displayed a perifascicular pattern and in IBM muscles they displayed a diffuse pattern (Online Resource 3).

#### Myofiber MHC-II expression regarding their myogenic status

The co-expression of MHC-II and CD56 by myofibers was assessed using serial muscle sections for every tested IIM muscle biopsy (n=69) (Figure 2, Table 2). A co-expression was found in 47 IIM patients (n=47/69, 68%) and was apportioned as follows: 10/22 for DM, 17/17 for IBM, 3/12 for IMNM, 7/7 for ASyS, and 10/11 for OM (Figure 2, Table 2). Of note, some muscle biopsies in which MHC-II immunostaining was negative had CD56 positive myofibers.

Fiber typing on serial muscle sections of IIM muscle biopsy with diffuse heterogeneous MHC-II myofiber immunostaining suggested that MHC-II positive myofibers were mainly type II myofibers (Online Resource 4).

### CAPILLARY IMMUNOHISTOCHEMISTRY STUDY

#### Capillary MHC-II abnormal immunostaining in IIM muscle biopsies

The MHC-II muscle biopsy immunostaining also identified capillary abnormalities (n=67/72, 93%), with quantitative (capillary dropout, n=32/64, 50%) and qualitative impairments (defined by architectural abnormalities, including dilated and leaky capillaries, n=65/72, 90%) (Figure 3, Table 3). Such qualitative impairments were frequently observed: leaky capillaries (n=48/62, 77%) and dilated capillaries (n=53/72, 74%) (Figure 3, Table 3).

**Fig. 3.**
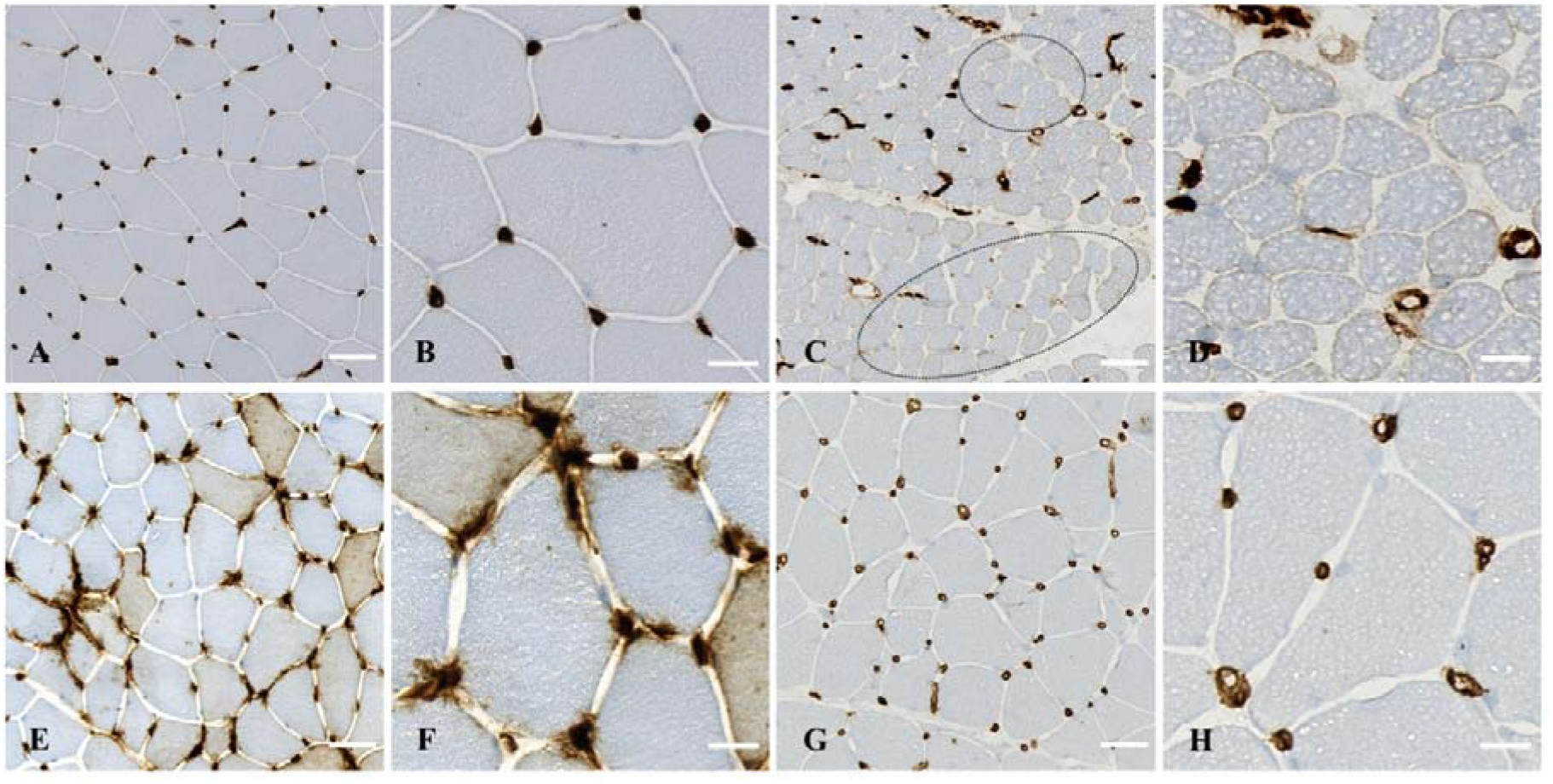
Capillary major histocompatibility complex (MHC) class II immunostaining in muscle biopsies from controls and IIM patients. (A,B) Normal MHC-II expression on capillaries. (C,D) Capillary dropout (dotted ellipses), (E,F) leaky capillaries, (G,H) dilated capillaries observable upon MHC-II immunostaining. Bars= 50μm (A,C,E) and 10μm (B,D,F).

#### Capillary MHC-II abnormal immunostaining differs according to IIM subgroup

While almost all the IIM subgroups presented MHC-II quantitative and qualitative capillary abnormalities, some subgroups displayed specificities (Table 3). MHC-II immunostaining identified that most IBM muscles presented almost no capillary dropout (1/9, 11%) but frequent dilated capillaries (n=15/17, 88%). In IMNM, capillary dropout was rare (2/12, 17%), while leaky and dilated capillaries were frequent (8/12, 67% for both). In comparison, DM, ASyS, and OM appeared to have high frequencies of quantitative and qualitative capillary lesions with the presence of both capillary dropout, leaky capillaries, and dilated capillaries (Table 3). CD31 was evaluated for 67 (93%) IIM patients and identified capillary dropout (n=39/67, 58%), leaky capillaries (46/67, 69%), and dilated capillaries (57/67, 85%). The concordance between the capillary abnormalities identified by MHC-II and CD31 immunostainings was high for both quantitative (capillary dropout, 83%) and qualitative impairments (leaky capillaries, 93% and dilated capillaries, 87%) (Figure 4, Table 3).

**Fig. 4.**
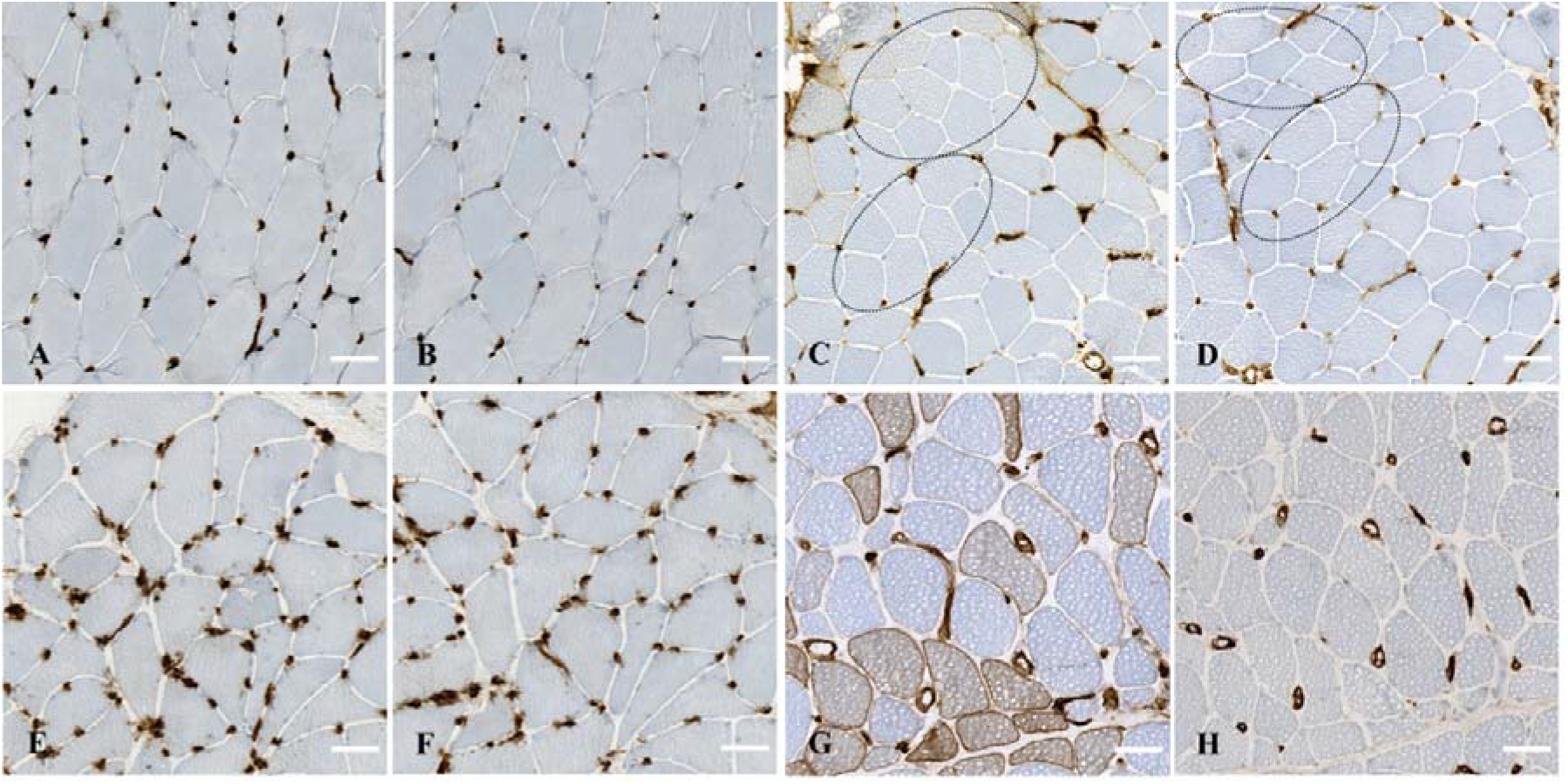
Capillary major histocompatibility complex (MHC) class II and CD31 immunostaining patterns in muscle biopsies from controls and IIM patients on muscle serial sections. Normal MHC-II (A) and corresponding normal CD31 (B) immunostaining. Capillary dropout observable upon MHC-II (C) and corresponding CD31 (D) immunostainings (dotted ellipses). Leaky capillaries observable upon MHC-II (E) and corresponding CD31 (F) immunostainings. Dilated capillaries observable upon MHC-II (G) and corresponding CD31 (H) immunostaining. Bars: 50μm (A,B), 100μm (C-H).

## Discussion

The present study identified the expression of MHC-II by myofibers as a potential diagnostic tool for IIM, providing elements for the distinction between IIM subgroups when taking into account the recent advances in IIM subgroup classification. The main patterns of myofiber MHC-II immunostaining found were: diffuse in IBM, perifascicular in ASyS, negative in IMNM, inconstantly positive in adult DM (mainly correlated with pediatric or neoplastic DM patients), and unspecific in OM (perifascicular, clustered, and diffuse heterogenous). Secondly, the analysis of MHC-II capillary immunostaining of IIM muscles identified quantitative and qualitative capillary abnormalities in IIM, with high frequencies and specificities according to IIM subgroups.

### Myofiber MHC-II immunostaining, a tool for the diagnosis of IIM subgroups

As progress is made regarding IIM subgroup delineation, and since related care differs according to subgroups, the need for specific biomarkers becomes more apparent [10]. Regarding MHC-II immunostaining in IIM muscle biopsies, most studies so far have evaluated MHC-II in polymyositis [6, 14, 17, 23, 24, 27, 33, 39, 43, 54], an entity which is slowly disappearing as patients are re-classified mainly as ASyS, IMNM, OM, and amyopathic DM [3, 28, 32]. Indeed, most previous studies were performed several decades ago, impeding the comparison with the currently used classification of IIM subgroups. Although few recent studies report interesting results, each of them analyzed only a particular subgroup [1, 4, 46], thus impeding the identification of MHC-II myofiber immunostaining variation between IIM subgroups.

Regarding IBM, the present study identified a mainly diffuse MHC-II myofiber immunostaining pattern, further strengthening and completing the results obtained in previous reports [14, 24, 43]. IBM diagnosis is complex [35], and many patients are currently facing diagnostic delays. The main reason for this is that IBM diagnosis is partly based on clinical features, notably distal muscle weakness, and on histological muscle lesions, notably rimmed vacuoles, which may appear late in the course of the disease [28]. Achieving a diagnosis has important clinical implications since IBM is the only IIM that do not benefit from immunosuppressive therapy. In that context, MHC-II immunostaining could be useful in reducing diagnostic delays.

Regarding the ASyS subgroup, the present results are in line with a recent study in which, among 33 ASyS and 17 DM patients, the authors identified a higher MHC-II positivity in ASyS compared to DM (81.8% vs 23.5%, respectively), and mainly with a perifascicular pattern [4].

Regarding IMNM, only a quarter of the tested biopsies showed MHC-II positive myofibers, a result in line with previously published cases [1, 36]. Another study however, reported a higher frequency of positivity in IMNM (43.7%) [43]. This could be explained by a difference in the diagnostic criteria applied, as herein none of the IMNM patients had anti-Jo1 or anti-PMScl antibodies.

In the DM subgroup, as reported in the literature, MHC-II positivity was more variable than in the other subgroups. In their study, Das et al. reported myofiber MHC-II positive immunostaining in 93% of DM cases [14], while in another study, MHC-II positive myofibers were present in 54.5% of DM cases [43], a result closer to the one obtained herein. The lack of documented autoantibodies and the perifascicular immunostaining found in all cases raise the question of whether ASyS cases were not considered as DM in the study by Das et al. [14]. Here, we noticed that the majority of DM patients with MHC-II positive myofibers were patients with either ongoing neoplasia or juvenile DM. Although this observation requires further investigation on larger groups of patients, the use of MHC-II immunostaining could be of great value for adult DM patients, as it could represent a new tool, easily usable in routine, to detect a paraneoplastic process. Regarding juvenile cases, in line with the present results, a recent study by Schänzer et al. identified a positive MHC-II myofiber immunostaining in 7 out of 9 cases of juvenile DM, with a mild intensity and a scattered distribution, except for 2 cases which showed a perifascicular pattern [46]. Another study, in 2009, found MHC-II positive myofiber immunostaining in only 28% of juvenile DM muscles [45]. Since that time however, progresses have been made in juvenile DM diagnostic criteria. Moreover, a large number of patients from that study had received treatment prior to their biopsy, and the age cut-off for defining juvenile DM differed from the study herein.

With regards to OM, the very variable patterns of MHC-II myofiber positivity may reflect the lack of specificity of this IIM subgroup and argue for the need to further refine their classification.

Overall, the present results show that MHC-II immunostaining could be a valuable diagnostic tool for IIM diagnosis and a useful parameter for distinguishing IIM subgroups.

### Input for the understanding of IIM pathogenesis

MHC-II molecules are known to play a pivotal role in the induction and regulation of immune responses through their ability to present antigens to CD4+ T lymphocytes. The expression of MHC-II by myofibers in some IIM subgroups question the potential immune role of myofibers. Indeed, while the expression of MHC-II molecules is constitutive of APCs (monocytes/macrophages, B cells, and dendritic cells), this expression appears to be inducible in most cell types and tissues under specific inflammatory conditions [30]. In the present work, the fact that myofibers express MHC-II raises the question of myofibers acting as potential APCs. Since the MHC-II positive myofibers also express CD56, it appears that the myofibers potentially acting as APCs are myofibers that are undergoing regeneration or are at least activated muscle cells. Some studies have demonstrated the potential role of myogenic cells as APCs [12, 53]. Another study has demonstrated that, for some IIM patients, the myofibers expressing MHC-II also express the intracellular adhesion molecule-1, a molecule required for the stabilization of the immunological synapse between MHC-peptide complex and T cells [6].These considerations advocate for an active role of myofibers in the complex dysimmune cascade that underlies IIM pathogenesis.

Another interesting point is the observation that type II myofibers (identified thanks to multiple techniques) were the myofibers that preferentially express MHC-II in case of heterogeneous positivity. This finding raises the hypothesis of a potential link between the glycolytic/oxidative metabolism of myofibers and their involvement in the immune process. Also, this observation is in line with the type II myofibers atrophy reported in myopathy associated with systemic inflammatory disorders [8].

Concerning underlying mechanisms, IFN-γ has been identified as a strong inducer of MHC-II cellular expression [30]. Interestingly, a recent study demonstrated that IBM muscles display a strong IFN type II (i.e., IFN-γ) signature which is not the case of other IIMs [41]. In this context, the mainly diffuse expression of MHC-II observed in IBM myofibers is in line with such a mechanism.

The identification of distinct patterns of MHC-II myofiber immunostaining among IIM subgroups also confirms the progress made in delineating this group of diseases and advocates for pathogenic-specific processes among these subgroups. For example, the mainly perifascicular MHC-II myofiber immunostaining found in ASyS strengthens the consideration of a specific disease, individualized from the OM subgroup. Regarding DM, the present findings support the idea of distinct pathogenic processes between juvenile and adult DM, which has been previously discussed in another study [46]. Indeed, while sharing pathognomonic cutaneous lesions and muscle inflammation, some clinical features differ between adult and pediatric cases, notably the TIF1γ -associated malignancy and anti-MDA5-associated rapidly progressive interstitial lung disease, which are found only in adult DM [50]. In the present study, the juvenile cases presenting MHC-II immunostaining all occurred in pre-puberty children while those with negative MHC-II immunostaining were adolescents. Moreover, in the adult paraneoplastic DM cases, the MHC-II negative cases were associated with ongoing ovarian adenocarcinoma, while those that showed MHC-II positive myofiber immunostaining were melanoma and bronchial adenocarcinoma cases. Altogether, this questions the hormonal implication during DM pathogenesis, and these results call for larger studies.

### Capillary impairment, a frequent element in all IIM subgroups

The present study identified abnormal muscle capillary immunostaining (by both MHC-II and CD31) as a common feature of IIMs. The observed microvasculature changes observed were both quantitative and qualitative impairments and found in various proportions in the different IIM subgroups. While capillary dropout appeared practically absent from IBM and IMNM muscles, qualitative abnormalities appeared to be largely shared by all IIM subgroups, a result in line with the only other study having evaluated capillary impairment in the currently defined IIM subgroups [16]. When assessing in more detail the types of abnormalities present, the current study identified both dilated and leaky capillaries. While dilated capillaries have rarely been described [16], these could resemble the pipestem capillaries reported by several other studies. These are defined by a thickening of the capillary walls, characterized by the absence of undulating tubules and the presence of an amorphous material other than amyloid [5, 25, 47]. To our knowledge, leaky capillaries have never been reported in IIMs. The observation of these capillaries abnormalities using both MHC-II and CD31 immunostaining argues for a non-artefactual result. In such capillaries, the sprout aspect emanating from endothelial cells resembles that described when pre-existing capillaries incorporate resident or circulating endothelial progenitor cells [21]. This result needs to be confirmed on larger cohorts and should be further investigated. In terms of diagnosis, CD31 immunostaining appeared more efficient to detect capillary dropout and dilated capillaries, while MHC-II immunostaining identified more efficiently leaky capillary lesions.

To date, microvasculature changes in IIMs have been mainly described in DM cases, and these include endothelial inclusion [11, 16, 18, 38], capillary depletion [7, 20, 37], and a significant increase in neovascularization, particularly in juvenile DM [34]. Interestingly, vasculopathy has been significantly linked to muscle damage [13], and to the severity of the disease, in both juvenile and adult DM [19, 20]. In line with the present findings, a recent study using ultrastructural analysis on 60 patients identified that capillary dropout was mainly present in DM, ASyS, and scleromyositis (an entity included in the OM subgroup) [16]. Regarding the OM subgroup, two recent studies identified basal membrane thickening and reduplications, endothelial activation, and pericyte proliferation in scleromyositis [16, 48]. One study using ultrastructural analysis reported a thickening of basal membrane in IMNM and basal membrane reduplication, increased number of pericyte processes, and endothelial activation in IBM [16], while another study reported that the microvascular architecture in IBM was distorted but without providing details [52].

Considering the potential pathogenic processes at play, one study demonstrated that polymyositis and DM were associated with phenotypic and functional dysregulation of endothelial precursor cells, which may be related to IL-18 and IFN-I [15].

### Limitation and perspectives

Although carried out on a small retrospective series, the present work allowed to evaluate MHC-II immunostaining in the 5 well-defined IIM subgroups. The blind pathological analysis was performed by 3 trained muscle pathologists, allowing an unbiased histological interpretation. The qualitative capillary impairments identified by MHC-II and CD31 immunostaining call for further investigation, notably by including an ultrastructural analysis. The mechanisms underlying such alterations also remain to be addressed.

## Conclusion

While recent expert opinion (EURO-NMD pathology working group) has recommended that MHC-II immunostaining of muscle biopsy remains optional, the present work demonstrates that MHC-II expression patterns allow to distinguish between several IIM subgroups. The present data thus argue for the inclusion of MHC-II immunostaining in the routine histological diagnosis of IIMs. Further investigations regarding capillary alterations and pathogenic mechanisms in IIMs are required.

## Data Availability

All data produced in the present study are available upon reasonable request to the authors

## Declarations

### Funding

MR was supported by the Ecole de l’Inserm Liliane Bettencourt Programme, LL by the Fondation pour la Recherche Médicale, and LG by AFM-Téléthon. The authors have no competing interests to declare that are relevant to the content of this article.

## Consent to participate and to publish

In accordance with the French legislation, informed consent was obtained from all individuals, and the study was validated by the local ethics committee and registered at the national data protection agency (Commission Nationale de l’Informatique et des Libertés; Approval # 22_5767).

## Notes

### Competing Interest Statement

The authors have declared no competing interest.

### Funding Statement

AFM-Telethon (number 23652, Laure GALLAY)

### Author Declarations

Ethics committee of Hospices Civils de Lyon, France, gave ethical approval (Approval # 22_5767) for this work

